# Study of association of migraine susceptibility genes with common migraine in 200,000 exome-sequenced UK Biobank participants

**DOI:** 10.1101/2021.12.02.21267184

**Authors:** Katherine Alexis Markel, David Curtis

## Abstract

**Background:** A number of genes have been implicated in rare familial syndromes which have migraine as part of their phenotype but these genes have not previously been implicated in the common form of migraine.

**Methods:** Among exome-sequenced participants in the UK Biobank we identified 7,194 migraine cases with the remaining 193,433 participants classified as controls. We investigated ten genes previously reported to be implicated in conditions with migraine as a prominent part of the phenotype and carried out gene and variant based tests for association.

**Results:** We found no evidence for association of these genes or variants with the common form of migraine seen in our subjects. In particular, a frameshift variant in *KCNK18*, F139Wfs*24, which had been shown to segregate with migraine with aura in a multiply affected pedigree was found in 196 (0.10%) controls as well as in 10 (0.14%) cases (χ^2^ = 0.96, 1 df, p = 0.33).

**Conclusions:** Since there is no other reported evidence to implicate *KCNK18*, we conclude that this gene and its product, TRESK, should no longer be regarded as being involved in migraine aetiology. Overall, we do not find that rare, functional variants in genes previously implicated to be involved in familial syndromes including migraine as part of the phenotype make a contribution to the commoner forms of migraine observed in this population.

## Introduction

Migraine is a disabling primary headache disorder which has a 1 year prevalence of about 12%, peaking in middle age and then reducing thereafter (Lipton *et al*., 2007). It is characterized by severe pain and is sometimes preceded by neurosensory aberrations known as aura, while associated symptoms include nausea, vomiting, photophobia, phonophobia, tiredness, irritability, reduced concentration, and problems with cognition (Goadsby *et al*., 2017; Sutherland, Albury and Griffiths, 2019). Migraine may have major socio-economic impacts on individuals’ quality of life, their families, and society due to their greater healthcare needs and reduced work productivity (Becker *et al*., 2007; Ferrari *et al*., 2015).

Migraine has been shown to have a strong genetic component, with heritability estimates ranging from 34-64%, with common genetic variants with small effect sizes contributing to risk of the common form of the disease while some rare monogenic conditions can include migraine as part of the phenotype (Honkasalo *et al*., 1995; Russell and Olesen, 1995; Mulder *et al*., 2003; Bron, Sutherland and Griffiths, 2021). A recent meta-analysis of genome-wide association studies (GWASs) of migraine identified 38 implicated loci (Gormley *et al*., 2016). Monogenic conditions with migraine as part of the phenotype include familial hemiplegic migraine (FHM), sporadic hemiplegic migraine (SHM), retinal vasculopathy with cerebral leukoencephalopathy and systemic manifestations (RVCL-S), familial advanced sleep-phase syndrome (FASPS), and cerebral autosomal dominant arteriopathy with subcortical infarcts and leukoencephalopathy (CADASIL). In FHM and SHM migraine is a core feature of the phenotype whereas in the other conditions migraine can appear before other symptoms emerge.

A number of ion-regulating genes have been implicated in FHM, which is a rare monogenic form of migraine with aura that also includes motor disturbances and hemiparalysis (Goadsby *et al*., 2017). These include *CACNA1A, ATP1A2, SCN1A*, and possibly *PRRT2*, though there is debate as to whether the latter is disease-causing or modifying (Ophoff *et al*., 1996; De Fusco *et al*., 2003; Dichgans *et al*., 2005; Riant *et al*., 2012; Pelzer *et al*., 2014). Variants which have been implicated in FHM differ in their predicted effects, with *ATP1A2* and *PRRT2* having loss-of-function (LOF) variants, *CACNA1A* having gain-of-function (GOF) variants, and *SCN1A* variants having a complex impact on protein function (Pietrobon and Moskowitz, 2013; Sutherland, Albury and Griffiths, 2019). Paroxysmal non-kinesigenic dyskinesia (PNKD) is an autosomal dominant episodic movement disorder caused by variants in the *PNKD* gene (Shen *et al*., 2011, 2015). There is a report of a family with PNKD accompanied by FHM associated with a frameshift variant in PNKD (Gardiner *et al*., 2015).

SHM is similar to FHM, except that it develops in the absence of a family history of hemiplegic migraine (Kovermann *et al*., 2017). Variants in *SLC1A3* have been identified in individuals with SHM and are thought to induce the migraine phenotype via a LOF mechanism that disrupts glutamate reuptake and leads to neuronal hyperexcitability (Jen *et al*., 2005; Kovermann *et al*., 2017; Sutherland, Albury and Griffiths, 2019; Paucar *et al*., 2020).

With respect to the other conditions, variants in *TREX1* have been implicated in RVCL-S (Richards *et al*., 2007; Stam *et al*., 2009), variants in *CSNK1D* in FASPS (Brennan *et al*., 2013) and variants in *NOTCH3* in CADASIL (Joutel *et al*., 1996).

Two previous studies have specifically looked at *NOTCH3* variants associated with CADASIL in UK Biobank participants. Masoli and colleagues studied the imputed data of 451,424 UKBB participants of European descent for two missense variants predicted to be pathogenic by VEP (Variant Effect Predictor), p.R1231C and p.A1020P, and found that individuals with the former variant had higher diastolic and systolic blood pressure and an increased rate of incident stroke, while individuals with the latter variant only had higher diastolic blood pressure (Masoli *et al*., 2019). Since almost all pathologic variants in *NOTCH3* involve alterations in the number of cysteines in the protein’s epidermal growth factor-like repeat domains, Rutten and colleagues examined these variants in 50,000 UKBB participants and found that these factors were associated with a very broad range of phenotypes from normal to CADASIL, with migraines often presenting decades before other symptoms (Rutten *et al*., 2020).

A single pedigree has been reported in which migraine with aura segregated with a frameshift variant (F139Wfs*24) in the *KCNK18* gene which encodes the TRESK two-pore potassium channel protein (Lafrenière *et al*., 2010). Subsequent studies suggested another frameshift variant (Y121Lfs*44) may impair protein function in a similar manner (Royal *et al*., 2019). These variants produce an additional start codon (ATG) within the reading frame, which may lead to the production of TRESK protein fragments that are thought to downregulate *KCNK18* expression via a feedback loop and also interfere with normal channel activity in a dominant negative fashion (Lafrenière *et al*., 2010; Lafrenière and Rouleau, 2011, 2012; Andres-Enguix *et al*., 2012; Royal *et al*., 2019).

We sought to investigate whether variants impacting the function of these ten genes might contribute to risk of developing commoner, less familial forms of migraine in the general population. In order to do this we investigated whether functional variants were associated with a clinically defined migraine phenotype in 200,000 exome-sequenced UK Biobank participants.

## Methods

The approach used was similar to that previously described for other phenotypes (Curtis, 2021). The UK Biobank consists of 500,000 volunteers who have undergone extensive phenotyping and who have provided biological samples. They are on average somewhat older and more healthy than the British population as a whole. Exome sequence data has been released for 200,627 participants and these variant call files were downloaded after genotype-calling by the UK Biobank Exome Sequencing Consortium using the GRCh38 assembly with coverage 20X at 95.6% of sites on average (Szustakowski *et al*., 2020). UK Biobank had obtained ethics approval from the North West Multi-Centre Research Ethics Committee which covers the UK (approval number: 11/NW/0382) and had obtained written informed consent from all participants. The UK Biobank approved an application for use of the data (ID 51119) and ethics approval for the analyses was obtained from the UCL Research Ethics Committee (11527/001). All variants were annotated using the standard software packages VEP, PolyPhen and SIFT (Kumar, Henikoff and Ng, 2009; Adzhubei, Jordan and Sunyaev, 2013; McLaren *et al*., 2016). As described previously, population principal components were obtained using version 2.0 of *plink* (https://www.cog-gemonics.org/plink/2.0/) with the *commands –maf 0*.*1 –pca 20 approx* (Chang *et al*., 2015; Galinsky *et al*., 2016; Curtis, 2021).

In order to define participants as cases we used a similar approach as that described previously and combined information about recorded diagnoses and about medication (Curtis, 2020a). Participants who had an ICD-10 diagnosis code of G43.* and/or who reported taking one of a number of medications which are relatively specifically indicated as treatments for migraine were classified as cases while the remainder were taken to be controls. The list of the medications used to define cases is provided in Supplementary Table S1.

Tests for association were carried out at the level of the gene, of categories of variant within each gene and at the level of individual variants. In order to test for association at the level of the gene, weighted burden analysis was carried out using the SCOREASSOC program (Curtis, 2012, 2019). For each variant a weight was assigned according to its predicted effect on gene function, with variants with a more severe impact being allocated a higher weight so that, for example, stop gained variants were assigned a weight of 100 while missense variants were assigned a weight of 5. The full set of variant types and weights is presented in Supplementary Table S2. Attention was restricted to rare variants with minor allele frequency (MAF) <= 0.01 in both cases and controls. As previously described, variants were also weighted by MAF so that variants with MAF=0.01 were given a weight of 1 while very rare variants with MAF close to zero were given a weight of 10 (Curtis, 2020b). For each variant the functional weight was multiplied by the frequency weight to produce an overall weight and then for each subject the weights of the variants carried by that subject were summed to produce a weighted burden score which was included in a logistic regression analysis with sex and 20 principal components as covariates. We have previously shown that this process adequately controls test statistic inflation in this population (Curtis, 2021). The statistical significance for association between migraine and the weighted burden score for each gene was summarised as a signed log p value (SLP), which is the log base 10 of the p value given a positive sign if the score is higher in cases and negative if it is higher in controls.

The variant types were also grouped into broader categories such as intronic, splice site, protein altering, as shown in Supplemental Table S2, in order to test whether any particular category of variant within a gene was associated with migraine risk. As described previously, logistic regression analyses were performed using the counts of the separate categories of variant as predictor variables, again including principal components and sex as covariates, to estimate the effect size for each category (Curtis, 2021). The odds ratios associated with each category were estimated along with their standard errors and the Wald statistic was used to obtain a p value, except for categories in which variants occurred fewer than 50 times in which case Fisher’s exact test was applied to the variant counts. The associated p value was converted to an SLP, again with the sign being positive if the mean count was higher in cases than controls.

We also examined specific missense variants which had previously been reported as implicated in the migraine-associated syndromes listed above. After a thorough literature review, we compiled a list of 222 such variants in the ten genes included for this study, as listed in Supplementary Table S3. For variants observed in 20 or more subjects, counts were compared between cases and controls. For rarer variants, the pooled counts for each gene were compared.

Additionally, because of the report implicating a *KCNK18* frameshift variant we compared the counts of all frameshift variants in this gene individually and collectively.

Data manipulation and statistical analyses were performed using GENEVARASSOC, SCOREASSOC and R (R Core Team, 2014).

## Results

Using the process described to define cases based on assigned ICD10 diagnosis and/or migraine specific medication, there were 7,194 cases, of whom 78.2% were female with mean age 55.2 (SD=7.9), and 193,433 controls, of whom 54.2% were female with mean age 56.5 (SD=8.1).

Gene-wise weighted burden analysis did not find evidence for association of rare, functional variants with migraine for any of the ten genes tested. Table 1 shows the SLP obtained for each gene of which the highest is 1.58 for *ATP1A2*, equivalent to *p*=0.026. This result is not statistically significant after correction for multiple testing and none of the SLPs for the remaining genes approaches significance.

**Table 1.** Results of weighted burden analysis testing for association with migraine. Statistical significance is expressed as the signed log *p* value (SLP), with positive values indicating that rate, functional variants in the gene are associated with increased risk of migraine.

| Gene | SLP |
| --- | --- |
| *ATP1A2* | 1.58 |
| *CACNA1A* | 0.33 |
| *CSNK1D* | -0.05 |
| *KCNK18* | 0.85 |
| *NOTCH3* | -0.17 |
| *PNKD* | 0.99 |
| *PRRT2* | -0.19 |
| *SCN1A* | -0.59 |
| *SLC1A3* | -0.37 |
| *TREX1* | -0.08 |

Likewise, there was no category of variants within any gene that produced significant evidence of association with the migraine phenotype. Detailed results are provided in Supplementary Table S4.

Of the individual missense variants identified from the literature, 13 were observed in 20 or more participants. Results for these variants are shown in Table 2 and it can be seen that none of them is markedly more common in cases than controls. E492K in *ATP1A2* occurs in 18 cases versus 298 controls, a results which is nominally significant at p=0.04 but again this is not statistically significant after correction for multiple testing. The cumulative totals for rarer variants in each gene are shown in Table 3 and again it can be seen that these are not commoner among cases.

**Table 2.** Counts of missense variants previously reported to be implicated in migraine phenotypes for variants observed in 20 or more participants.

| Gene (canonical transcript) | Amino Acid Change | Control carriers (frequency) | Case carriers (frequency) |
| --- | --- | --- | --- |
| *CACNA1A* (ENST00000638029) | E1018K | 2442 (0.01262) | 88 (0.01223) |
| *ATP1A2* (ENST00000361216) | Y9N | 1465 (0.00757) | 58 (0.00806) |
|  | R65W | 33 (0.00017) | 2 (0.00028) |
|  | E492K | 298 (0.00154) | 18 (0.00250) |
| *SCN1A* (ENST00000303395) | T1174S | 438 (0.00226) | 14 (0.00195) |
|  | R1928G | 684 (0.00354) | 18 (0.00250) |
| *PRRT2* (ENST00000567659) | E23K | 1041 (0.00538) | 25 (0.00348) |
|  | L372F | 62 (0.00032) | 2 (0.00028) |
| *NOTCH3* (ENST00000263388) | R1143C | 67 (0.00035) | 1 (0.00014) |
|  | R1201C | 19 (0.00010) | 1 (0.00014) |
|  | C1222G | 82 (0.00042) | 3 (0.00042) |
|  | R1231C | 116 (0.00060) | 5 (0.00070) |
| *SLC1A3* (ENST00000265113) | R499Q | 50 (0.00026) | 3 (0.00042) |

**Table 3.** Cumulative counts of missense variants previously reported to be implicated in migraine phenotypes for variants observed in fewer than 20 participants.

| Gene (canonical transcript) | Control carriers<br>(frequency) | Case carriers<br>(frequency) |
| --- | --- | --- |
| CACNA1A (ENST00000638029) | 13 (0.00007) | 1 (0.00014) |
| ATP1A2 (ENST00000361216) | 5 (0.00003) | 3 (0.00042) |
| SCN1A (ENST00000303395) | 0 (0.00000) | 0 (0.00000) |
| PRRT2 (ENST00000567659) | 6 (0.00003) | 0 (0.00000) |
| CSNK1D (ENST00000398519) | 0 (0.00000) | 0 (0.00000) |
| NOTCH3 (ENST00000263388) | 111 (0.00057) | 6 (0.00083) |
| SLC1A3 (ENST00000265113) | 35 (0.00018) | 0 (0.00000) |

The frameshift variant in *KCNK18* which had been reported to segregate with migraine with aura, F139Wfs*24, was found in 196 (0.10%) controls and 10 (0.14%) cases (χ2 = 0.96, p = 0.33). The Y121Lfs*44 frameshift variant was found in 205 (0.11%) controls and 6 (0.08%) cases (χ2 = 0.34, p = 0.56). We identified an additional 20 controls and 0 additional cases with other frameshift variants in *KCNK18*, bringing the overall numbers to 421 (0.22%) controls and 16 (0.22%) cases (χ2 = 0.01, p = 0.93).

## Discussion

Overall, we do not observe an association between migraine risk and an excess of rare variants predicted to impact function in any of the ten genes previously implicated in rare familial disorders with migraine as part of the phenotype. This applies to the three canonical FHM genes (*CACNA1A, ATP1A2*, and *SCN1A*) and a potential fourth FHM gene *(PRRT2*), as well as the other genes reported to cause syndromes in which migraine may be a feature. These findings contrast with those we obtained from a similar study of hyperlipidaemia in the same UK Biobank sample, in which we found that for genes known to cause rare, severe forms of familial hypercholesterolaemia there were very rare variants with large effects on risk but also less rare variants which made a wider contribution to hyperlipidaemia in the general population (Curtis, 2021).

Even when we focus on specific variants previously reported to be associated with migraine syndromes we do not observe a significant excess among our cases. Of course, the method we have used to define caseness is not as rigorous as could be applied in a specially designed study and the control samples will include some participants with migraine who have not been assigned a formal diagnosis and who are not taking a migraine-specific medication. Nevertheless, it is also possible that not all of the previously reported variants do in fact raise migraine risk.

Our findings for *KCNK18* are particularly noteworthy. The only evidence to support the claim that this gene has an aetiological role in migraine is the observation that a frameshift variant, F139Wfs*24, perfectly segregated with the phenotype of migraine with aura in a single large pedigree (Lafrenière *et al*., 2010). It has been the subject of several functional studies, most of which were conducted on cultured cells or in animal models (Lafrenière *et al*., 2010; Lafrenière and Rouleau, 2011; Andres-Enguix *et al*., 2012; Liu *et al*., 2013; Guo *et al*., 2014, 2019; Royal *et al*., 2019). However we cannot find any report that this variant or other functional variants in *KCNK18* have been observed in other migraine patients. We observe this specific variant in 196 controls as well as in 10 cases and overall frameshift variants in this gene occur at equal frequency in controls and cases. These findings are not compatible with the notion that F139Wfs*24 causes a very rare quasi-mendelian form of migraine and hence we conclude that *KCNK18* and its product TRESK should no longer be regarded as influencing migraine susceptibility. We note that the haplotype segregating with migraine in the original pedigree extends over 10 megabases and harbours dozens of genes. While we can appreciate that a frameshift variant would have appeared to be a plausible candidate, our new results rule out a role for this variant and we speculate that perhaps another gene within this haplotype is actually responsible for the migraine cases observed in the pedigree.

Using an exome-sequenced sample of 200,000 participants we have been unable to identify a role for previously identified genes implicated in rare familial disorders as contributing to the risk of the more common migraine phenotype. Sequence data for the remaining 300,000 participants will be made available in due course and with the increased sample size it will become feasible to systematically study all genes in order to attempt to identify some which do influence migraine risk.

## Data Availability

Raw data is available on application from UK Biobank. Relevant derived variables will be deposited in UK Biobank.

https://www.ukbiobank.ac.uk/

## Acknowledgements

The authors wish to acknowledge the staff of University College London’s Computer Science Department and their High Performance Computing Cluster. We would also like to thank the participants of the UK Biobank study who provided the data for this study. This work was carried out in part using resources provided by BBSRC equipment grant BB/R01356X/1. KM would like to thank The Migraine Trust for awarding her a Susan Haydon Bursary.

## Contributions

DC conceived the methods, developed the programs, and contributed to the analysis and writing of this paper. KM helped adapt the methods for the migraine phenotype, selected the genes and variants included in the study, contributed to the analysis of the data, and wrote the paper.

## Data Availability Statment

Raw data is available on application from UK Biobank. Relevant derived variables will be deposited in UK Biobank. The scripts and software used to carry out the analyses are available at: https://github.com/davenomiddlenamecurtis.

